# Asthma prevalence in southern Spain

**DOI:** 10.1101/2025.03.24.25324505

**Authors:** Alberto Moreno-Conde, Claudia Rodríguez-Vegas, Jesús Moreno-Conde, Pedro Guardia-Martínez, Angel Vilches-Arenas, Virginia De Luque-Piñana

## Abstract

**Background and objectives:** Asthma is a chronic respiratory condition with varying prevalence across different regions and populations. In Andalusia, there is lack of detailed data on the characteristics of asthma patients, limiting effective clinical management and healthcare planning. This study aims to characterize asthma-diagnosed patients in Andalusia.

**Materials and methods:** To our knowledge this study represents the largest cohort of asthma patients in Spain. We characterized 726006 asthmatic patients based on prevalence, exacerbations, comorbidities and pharmacological treatment. We covered the overall population with sub-analyses based on rural and urban distinctions and geographical differences.

**Results:** The overall asthma prevalence in Andalusia was 8,57%. The disease was more prevalent in females, although it was more common in males until the age of 15. The most frequent comorbidities was arthrosis, spondylosis. 70,73% of newly diagnosed asthma patients received treatment. For those who did SABA (31,56%), LTRA (16,13%) and systemic glucocorticoids (13,94%) were the most common therapies. After initial treatment, 35,15% of patients did not receive follow-up care, 28,68% stepped-up, 20,75% stepped-down and 15,43% switched treatments.

**Conclusions:** The obtained results expect to provide a detailed characterization of the Andalusian population including prevalence, comorbidities and treatment trajectories aligned with other Spanish regions. In addition, there were found geographical discrepancies with regard to asthma prevalence when comparing rural-urban settings and coastal-inland regions.

## 1. INTRODUCTION

Asthma is a chronic respiratory disease that affects individuals of all ages. The World Health Organization (WHO) currently estimates 300 million patients with bronchial asthma in the world. Prevalence shows enormous variability between different geographical areas, both in adults (between 0,2% and 21%) and in children (between 2,8% and 37,6%)^1^. Differences in asthma prevalence between different studies may be due to a wide variety of causes, including different exposures to risk factors, different diagnostic criteria, ethnic, geographic, socioeconomic, climatic or environmental variations^2^.

Asthma treatment is organized via primary and specialized care based on patient’s symptoms severity, factors surrounding the patient and characteristics of the disease itself^3^. To support clinicians in the management of patients with asthma, several national and international guidelines have been developed, which are frequently updated based on the latest research and insights. These guidelines suggest a stepwise treatment approach where treatment is initiated and tailored to evolution and control (symptoms, severity, disease control and future risk) of the individual patient^4^. The main objective of asthma treatment is to control the disease, prevent exacerbations, fixed airflow obstruction, and minimize mortality. Thus, a comprehensive, long-term and individualized strategy based on optimal and customed treatment is necessary ^5^.

Climatic conditions such as high humidity and temperature fluctuations can exacerbate asthma symptoms by affecting air quality and the dispersion of airborne allergens^6^. Most studies on the effects of pollen have focused on acute effects, as represented in Emergency Department Visits or hospital admissions, occasionally describing inconsistent results, or geographical discrepancies^7^. In regions like southern Spain, where pollen from trees, grasses and weeds is abundant, the risk of asthma exacerbations can increase significantly, particularly during peak pollen seasons.

The primary objective of this study is to comprehensively characterize asthma patients in Andalusia, evaluating the progression of asthma diagnosed patients over 17 years. Thus, demographic data, clinical profiles such as comorbidities and environmental exposures are analyzed. To expand knowledge on how patients with asthma are treated in the real world, we analyzed pharmacological dispensations. Our study explores asthma trends across different regions, focusing first on a general analysis, then distinguishing between rural and urban populations, and finally breaking down the data by region. Our goal with this study is to explore hypotheses that could lead to further follow-up research to address current gaps in clinical practice.

## 2. MATERIAL AND METHODS

The cohort includes patients with asthma diagnosis based on the physician’s judgment, which has been recorded and coded accordingly in the medical history (ICD classification) from 1 January 2005 to 31 December 2021. Patients entered the cohort on the date of diagnosis (index date). Records with errors were excluded from the analysis, this included cases with coding inaccuracies or missing data. Specifically, entries where the residence was outside Andalusia, diagnosis dates were inconsistent, deaths occurred before the observation period ended, or there were discrepancies related to the recorded sex of individuals, were removed. Patient characteristics were captured, we considered covariates such as patient’s residence, age and sex. Charlson Comorbidity Index was calculated and specific comorbidities of interest were taken into account. Variables considered for the study are shown in table 1.

**Table 1:** Variables taken into account for the study

| Data Type | Variables | Coverage Period |
| --- | --- | --- |
| Personal information | Patient's residence, age and sex | 2005 - 2021 |
| Health data | Asthma diagnosis date and comorbidities | 2005 - 2021 |
| Emergency department visits | Emergency department visit date | 2009 - 2021 |
| Drug dispensations | Drug dispensation records | 2005 - 2021 |

### Location

This was a retrospective study, carried out in Andalusia, with over 8 million inhabitants (exactly 8472407 in 2021). It is divided into eight provinces: Almeria, Cadiz, Cordoba, Granada, Huelva, Jaen, Malaga, and Seville.

### Personal information and health data

Clinical information was obtained from Diraya (Electronic Health Record System of Andalusia). This infrastructure includes data on the geolocation of the residence of the 8 million patients it covers. For the emergency department visits, we focused on asthma-related emergency visits according to International Classification of Disease (ICD) codes. Specifically, we utilized information on the patient’s residence, age, sex, comorbidities, date of emergency visit and drug dispensations.

### Analysis

This study is characterizing asthma diagnosed patients and thus is descriptive in nature. No statistical comparisons were performed. It was conducted using Python.

We analyzed asthma prevalence and exacerbations, using sex, age and comorbidities as covariates and calculating frequencies. An exacerbation was defined as an Emergency Department Visit. We studied the periodicity of this time series data in 2009–2021. Following normalization, the Fourier Transform was applied to decompose the time series into its frequency components, revealing the strength of different frequency components in the data. By examining the resulting frequency spectrum, the most significant periodic pattern is found.

For all patients, we investigated whether they had any drug dispensation prescribed for treating asthma (both maintenance and rescue therapy) during follow-up. For those who did, we studied the treatment trajectory, which is defined as the sequence of the respective respiratory drug classes over time. If a patient dispensed more than a drug class at the same time, it was considered as combination therapy. After constructing treatment trajectories for each patient, we counted the number of patients with the same treatment trajectory. Aggregated results (for trajectories that occurred in at least 0,5% of the population) are presented in the form of sunburst plots. Treatment switching and step-up/step-down treatment were also investigated. This part of the methodology is aligned with other papers^8^.

After assessing general asthma trends, we further analyzed asthma trajectories across different regions (coastal and inland) of Andalusia and across urban and rural areas.

## 3. RESULTS

Andalusia, is the largest and most populous autonomous community in Spain (8472407 inhabitants in 2021). Between 2005 and 2021, 725948 people were diagnosed with asthma, the prevalence of asthma in the studied population is approximately 8,57%, translating to 8568 cases per 100000 individuals. Geographically, the highest diagnosis rate per province inhabitants were found in Granada and Jaen with a prevalence of approximately 10% (annex table A1.1). As shown in table 2; 44,45% of the diagnosed patients correspond with individuals under the age of 20. Women had significantly higher asthma prevalence than men. Figure 1 shows that both in men and women, the prevalence of medical diagnosis of asthma reached its peak at 1 years old. Moreover, the prevalence was higher for males than for females until approximately the age of 15; however, this tendency reverses after that age, with women having higher prevalence than men overall (54%).

**Table 2:** Frequencies and proportions of patients diagnosed with asthma from 2005-2021 and with emergency room visits from 2009-2021.

|  | Asthma diagnoses (2005-2021) |  |  | Asthma exacerbations (2009-2021) |  |  |
| --- | --- | --- | --- | --- | --- | --- |
|  | N | % | % Province | N | % | % Province |
| **SEX** |  |  |  |  |  |  |
| Men | 332958 | 45,87% | - | 45951 | 41,46% | - |
| Women | 392990 | 54,13% | - | 64868 | 58,54% | - |
| **PROVINCE** |  |  |  |  |  |  |
| Almería | 55655 | 7,67% | 7,61% | 11164 | 10,07% | 1,53% |
| Cádiz | 111340 | 15,34% | 8,94% | 17504 | 15,80% | 1,40% |
| Córdoba | 69367 | 9,56% | 8,93% | 8047 | 7,26% | 1,04% |
| Granada | 96668 | 13,32% | 10,49% | 22420 | 20,23% | 2,43% |
| Huelva | 39277 | 5,41% | 7,47% | 4065 | 3,67% | 0,77% |
| Jaén | 61115 | 8,42% | 9,74% | 8463 | 7,64% | 1,35% |
| Málaga | 125464 | 17,28% | 7,40% | 17322 | 15,63% | 1,02% |
| Sevilla | 167062 | 23,01% | 8,58% | 21834 | 19,70% | 1,12% |
| **AGE** |  |  |  |  |  |  |
| [0,19] | 322670 | 44,45% | - | 48221 | 43,51% | - |
| [20,39] | 184806 | 25,46% | - | 29120 | 26,28% | - |
| [40,59] | 136923 | 18,86% | - | 20844 | 18,81% | - |
| [60,79] | 73359 | 10,11% | - | 11407 | 10,29% | - |
| >= 80 | 8190 | 1,13% | - | 1227 | 1,11% | - |

**Figure 1:**
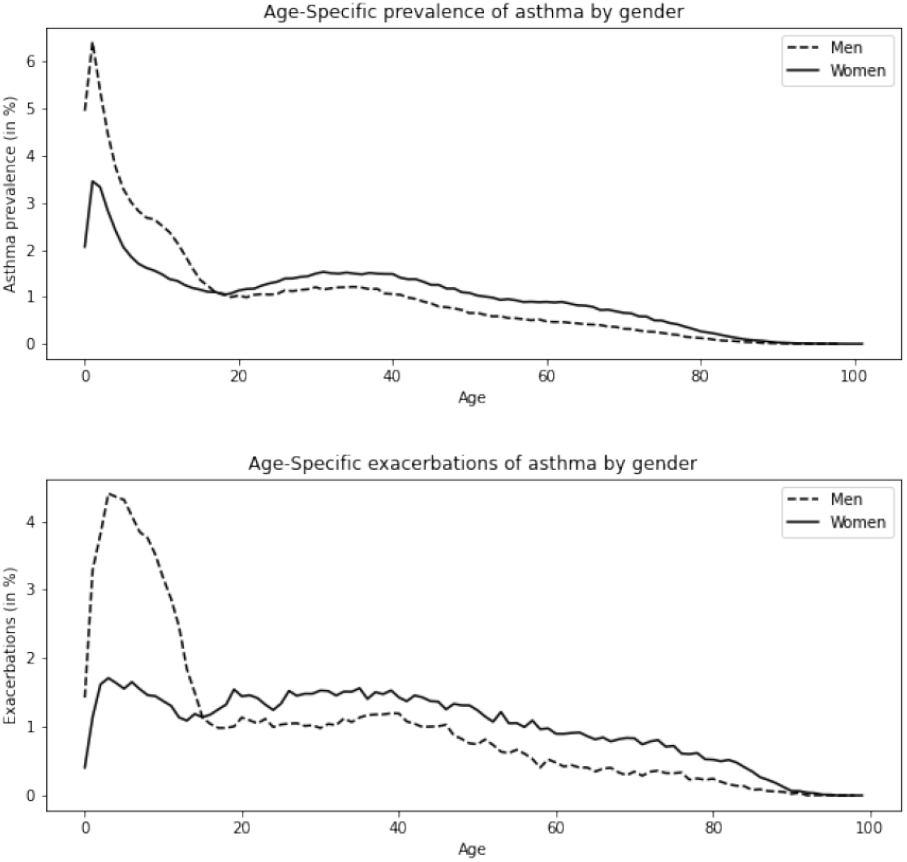
Gender and age-based comparison of asthma prevalence (2005 - 2021) and exacerbations (2009 - 2021). Two distinct lines are plotted to show the trends for men and women, allowing for a gender-based comparison of asthma prevalence across various ages.

Between 2009 and 2021, 15% of the considered cohort suffered an exacerbation (thus and emergency department visit). Mean daily emergency cases in Andalusia was 23,34 cases per day, furthermore during this period, only two days had no reported cases. Notably, the highest number of cases occurred on May 15, 2013, with a total of 232 cases. As shown in table 2; women had significantly more exacerbations than men. Figure 1 shows that the number of exacerbations was higher for males than for females until approximately the age of 15; however, this tendency reverses after that age, with women having higher prevalence than men overall (59%).

The time series corresponding to emergency visit series follows a periodic pattern. After normalizing the data, the dominant frequency of the Fourier Transform was found to be 182,61 days, thus the emergency data followed a repeating pattern approximately every 6 months (annex A1.2). We conducted an exploratory analysis grouping the data by month, and we added monthly mean environmental factors. Both in terms of diagnoses and emergency cases, May was the month with the highest number of reported cases, 19% of the exacerbations correspond with the peak concentration of grasses and olive trees.

Before diagnosis arthrosis, spondylosis (8%), dyslipidemia (5%) and atopic dermatitis (3%) were the most frequent non-respiratory comorbidities (annex table A1.3).

The sunburst plot for the treatment trajectories of asthma diagnosed patients is presented in figure 2. In the overall cohort, 70,73% of asthma diagnosed cases were treated (we considered treated patients as those who have at least one drug dispensation for asthma after diagnosis date). For those patients the five most popular lines of therapy were SABA - short-acting β2 antagonists (31,56%), LTRA - leukotriene receptor antagonists (16,13%), Systemic Glucocorticoids (13,94%), ICS + SABA combination therapy (8,86%) and ICS (7,87%). After first treatment, 35,15% of the patients did not receive any follow-up treatment. Furthermore, 28,68% of the patients stepped-up, 20,75% stepped down and 15,43% switched (more information in the annex section A1.4). Main results of treatment trajectories are shown in a sunburst graph (Figure 2).

**Figure 2:**
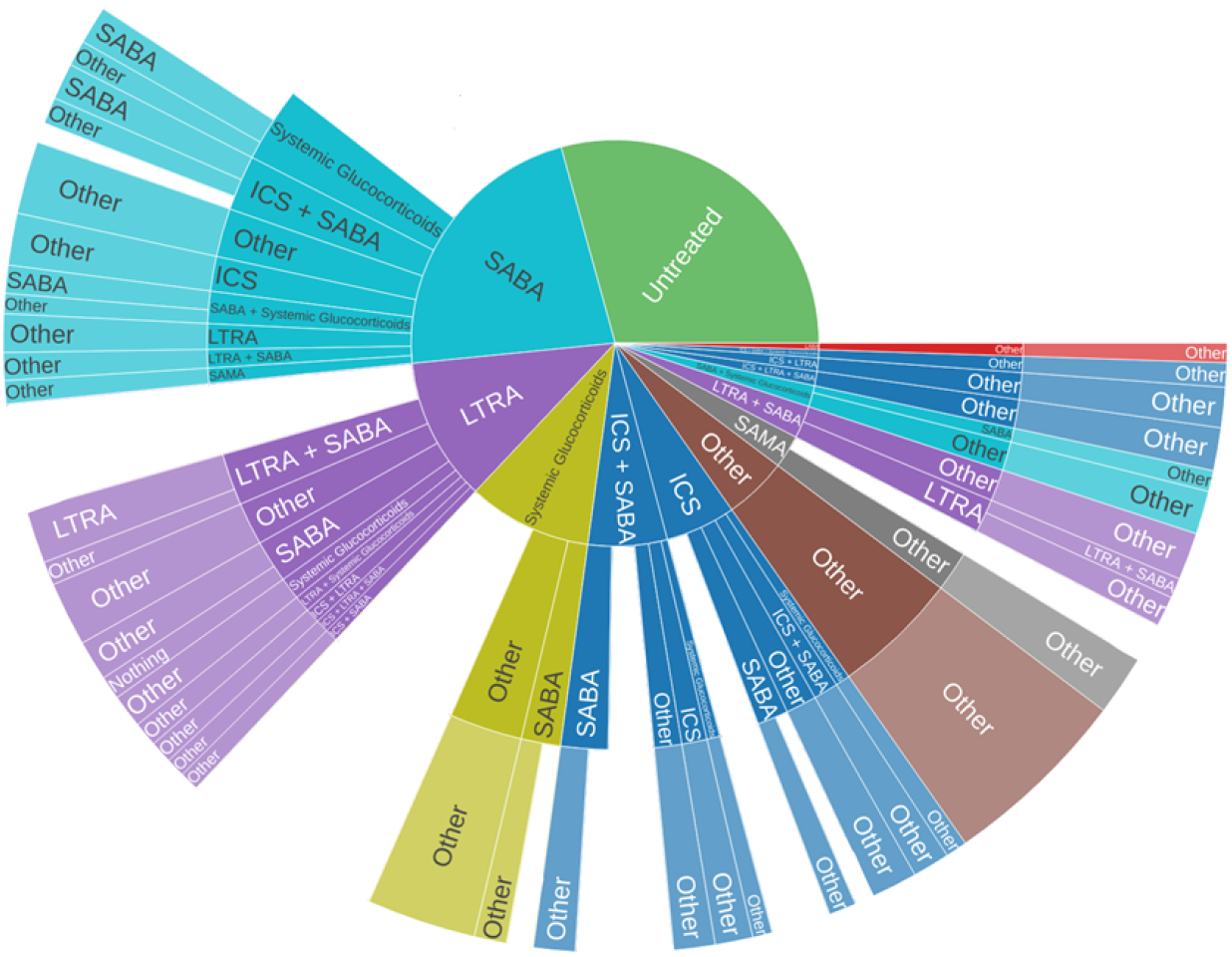
Sunburst plot of asthma treated patients, showing their first respiratory treatment in the center and subsequent pharmacological treatments in the surrounding layers. Each color represents the first treatment. Other corresponds to trajectories that occur in at less than 0,5% of the cases. ICS - inhaled corticosteroids; LABA - long acting β2 antagonists; LAMA - long acting muscarinic antagonist; LTRA - Leukotriene receptor antagonists; SABA - short-acting β2 antagonists; SAMA - short-acting muscarinic antagonist; S.G. - Systemic Glucocorticoids; PDE4 - phosphodiesterase.

After assessing the general asthma trends, we further analyzed asthma trajectories by taking into account demographic, socioeconomic, and environmental factors. Thus, we repeated the study across urban and rural areas (annex A1.5). In addition, we analyzed inland and coastal regions (annex A1.6).

In 2021, approximately 42,09% of the population resided in rural areas, while the majority, 57,91%, lived in urban regions. In rural areas, the prevalence of asthma was notably higher (11,40%). Conversely, urban areas reported a lower asthma prevalence (6,51%). However, the frequency of exacerbations of asthma remained similar between rural (15,10%) and urban areas (16,88%). Access to and coverage of asthma treatment showed a slightly difference, in rural areas, 72,18% received treatment and in urban areas, 68,89%. Comorbidities were higher in rural settings and treatment trajectories showed a similar distribution.

We classified Almeria, Cadiz, Huelva and Malaga as coastal region and Cordoba, Granada, Sevilla and Jaen as inland. In 2021, population was evenly distributed approximately 49,56% of the population resided in coastal areas, and 50,44%, lived in inland regions. Inland provinces showed greater asthma prevalence (9,23%) compared to coastal provinces(7,90%). However, the frequency of exacerbations of asthma remained similar between coastal (15,73%) and inland areas (16,01%). Access to and coverage of asthma treatment showed a slightly difference, in inland areas, 72,46% received treatment and in coastal areas, 68,68%. Comorbidities were higher in inland settings and treatment trajectories showed a similar distribution. In inland regions, the majority of exacerbations occurred in May, while in coastal areas, exacerbations were evenly distributed across the winter months without a clear peak. Interestingly, even with this increase in May, the exacerbation rates did not show significant differences.

Furthermore, we compared pandemic (2020-2021) and pre-pandemic (2018-2019) years. In terms of diagnoses, non-COVID years recorded an average of 59201 diagnoses of asthma. During the COVID years, this average dropped to 42590 diagnoses per year (28,06% decrease). In terms of exacerbations, in pre-COVID years, the annual average of visits to emergency departments was 22191 visits per years. However, during the COVID years, this number significantly dropped to an average of 15565 visits per year (29,86% decrease).

## 4. DISCUSSION

In our population, the prevalence remains within the described range (8,57%) matching the prevalence described in other works^2^. Other population characteristics such as average age (27,35 years) and gender distribution (54% female patients, 46% male) also align with what is described in the referenced literature^9^.

We included patients diagnosed with asthma by a physician from 0 years old. This approach, while aligned with current research practices, may include instances where asthma has not been definitively confirmed, reflecting the complexities of early childhood respiratory pathology^10^. This could be considered as a limitation, since patients under 3 years old with recurrent wheezing may not develop asthma later on. However, many asthma cases present symptoms in the early years of life, and it is the most prevalent chronic disease in childhood, with a prevalence of 10%. In our study, the prevalence in children under 6 years of age is 24,22%, which may be overdiagnosing the pathology due o the difficulties in diagnosis at early ages ^11^.

According to our results, the prevalence was higher for males than for females until approximately the age of 15; however, this tendency reverses after that age, with women having higher prevalence than men overall (54%). Interestingly, similar findings were observed in a study conducted in Catalonia, where the reversal in asthma prevalence occurred later, around the age of 30^12^. These consistent trends across different populations highlight a shift in asthma risk that emerges during adulthood, particularly among women.

Comorbidities may complicate exacerbations, disease management and control of the asthma patient’s symptoms. Conditions such as rhinitis, chronic rhinosinusitis, gastroesophageal reflux disease (GERD), obesity, obstructive sleep apnea, atopic dermatitis, food allergies, depression, and anxiety are often associated with asthma^13^. In this line of research in our cohort, the most prevalent comorbidities included anxiety disorder (1%), atopic dermatitis (3%) and obesity (1%).

Regarding the absence of rhinitis as a comorbidity in this large cohort of patients, it may seem that the importance of rhinitis as a disease, either individually or associated with asthma and/or conjunctivitis, could be underestimated due to the severity of another condition in the patient, which may result in rhinitis not being reported despite its presence ^14^. It should be taken into account that we considered comorbidities previous to diagnosis, thus the low percentages (in comorbidities in general) can be explained by the fact that these conditions have not yet developed, but may potentially emerge in the future.

The diagnoses and exacerbations peak in May could be associated with pollen levels, since it is known that pollen has an influence on asthma exacerbations^15^. In our geographic area, in inland provinces, the highest pollen levels are recorded in May, mainly related to grasses (gramineae) and olea.

We encountered several geographical discrepancies with regard to asthma prevalence when comparing rural-urban settings and coastal-inland regions of Andalusia. Several studies focusing on children have reported a lower prevalence of asthma in rural regions when compared to urban areas^16,17^. In our cohort (covering both children and adults) we found out that despite the higher prevalence of asthma in rural areas (11,40%), patients living in urban areas were more likely to experience an exacerbation (16,88%). Inland provinces showed greater asthma prevalence (9,23%) compared to coastal provinces (7,90%). Interestingly, in inland regions the majority of exacerbations occurred in May (13,75%), possibly due to increased pollen and other allergens prevalent during spring. However, in coastal areas, asthma exacerbations tend to be more evenly distributed across the winter months (average of 7,84% from December to February). This could be attributed to the relatively milder and more stable coastal temperatures, which prevent sudden, freezing events that are known triggers of asthma exacerbations^18^.

The analysis between pandemic and pre-pandemic years shows that the pandemic had a significant impact on diagnosis and emergency department visits, with a significant drop in both, as expected^19^. Public health responses during the COVID-19 crisis, including social isolation and mask usage, may have helped curb the circulation of common seasonal viruses, while reduced transport activity led to a drop in pollution levels.

In our cohort 70,73% patient diagnosed with asthma were treated. As expected according to literature most adults with asthma initiated treatment with SABA monotherapy (31,56% of first-line treatments)^20, 21^. This was followed by leukotriene receptor antagonists (LTRA) (16,13%) and inhaled corticosteroids (ICS) (13,94%). This results are aligned with those reported in Europe^8^. Regarding treatment switching and step-up/step-down therapy, we observed both increases and decreases in treatment among patients with asthma, which is also consistent with existing literature. Along with the need for regular follow-up and adjustments tailored to asthma patients, it suggests that pharmacological treatments may be customized to the individual needs of patients, taking into account factors such as symptoms, severity, disease control, and future risk. In this regard, it is necessary to reinforce asthma patients’ treatment according to season and location. It’s important to note that despite changes in guidelines during the study period, the treatment trajectories observed remained consistent over the years.

A similar study from Catalonia reported that SABAs were the most prescribed medication overall (62,6%), followed by systemic corticosteroids (43,3%) and combination therapies (ICS and bronchodilators, 41,9%). The Catalonian study also emphasized high prescription rates for systemic corticosteroids and bronchodilators. The differences in treatment patterns, particularly regarding establishing referral criteria and improving continuity of care systemic corticosteroids and combination therapies, may reflect variations in healthcare access, socioeconomic factors, and regional clinical practices.

Traditionally, consensus guidelines on asthma management established short-acting beta-agonists (SABA) as the first line of treatment. However, in recent revisions, inhaled corticosteroids, combined with bronchodilators (LABA), have been incorporated into the early therapeutic steps^5^. In this regard, our cohort did not show the influence of this new trend in asthma patient management. Therefore, a key objective for the care should be to update their treatments in accordance with the latest consensus guidelines.

To our knowledge, our study represents the largest cohort of asthma patients in Spain to date with a cohort of 725948 individuals. This increase in sample size enhances the ability to generalize findings and identify trends that might be missed in smaller studies.

This study offers an extensive characterization of real-world adult populations with asthma. It is important to acknowledge that the analysis of treatment patterns using observational data is constrained by the availability of drug dispensing information, which provides no information about the physician’s prescribed treatment plan or the patient’s adherence to it. To enhance clinical practice, it is crucial to examine discrepancies between (inter)national asthma guidelines and medication use in real-world settings, which could provide a better understanding of the adherence (or lack thereof) to these guidelines. Further research is needed to explore how treatment patterns evolve over time, especially in response to new guideline recommendations. With current treatments available, most patients could achieve control of their pathology, however for this to be possible, both good treatment adherence by the patient and better coordination among all professionals involved in the care of asthma patients (establishing referral criteria and improving continuity of care) are needed.

## Ethical approval

This research has been approved by the Andalusian Biomedical Research Ethics Committee Coordinator under protocol number 2403-N-20. The committee granted exemption from informed consent for retrospective studies based on the safeguard mechanisms defined in Spanish Law 3/2018 on personal data protection and digital rights, which mandates technical and functional separation between researchers and technicians responsible for patient data pseudonymization.

## Data Availability

All data produced in the present study are available upon approval from the Andalusian Health Service Access Committee.

## Abbreviations and acronyms

AEMET: State Meteorological Agency AEMET
WHO: The World Health Organization.

